# Twenty-site HDSS mapping after thoracoscopic sympathectomy for primary palmar hyperhidrosis: a secondary complete-case analysis of a multicenter randomized trial

**DOI:** 10.64898/2026.08.02.26359508

**Authors:** Nelson Wolosker, Niura Noro Hamilton, Miguel Lia Tedde, Marina Borri Wolosker, Wolfgang W Schmidt Aguiar, Hylas Paiva da Costa Ferreira, Fernando Luiz Westphal, Alexandre M Rodrigues Lima, Humberto A de Oliveira, Sergio Tadeu L F Pereira, Fabio de Oliviera Riuto, Guilherme C Resende, Marina Maria Krum Brenner, Daniel de Oliveira Bonomi, Caroline E Brero Valero, Paulo M Pego-Fernandes

**Affiliations:** Hospital das Clinicas, University of Sao Paulo, Sao Paulo, Brazil; Faculdade Israelita de Ciencias da Saude Albert Einstein, Sao Paulo, Brazil; Heart Institute (InCor), Hospital das Clinicas, University of Sao Paulo, Sao Paulo, Brazil; Hospital Universitario Oswaldo Cruz, Recife, PE, Brazil; Hospital Liga Norte Riograndense Contra o Cancer, Natal, RN, Brazil; Hospital da Universidade Federal do Amazonas, Manaus, AM, Brazil; Hospital Geral Dr. Cesar Cals, Fortaleza, CE, Brazil; Hospital de Base, Brasilia, DF, Brazil; Hospital Santa Isabel, Salvador, BA, Brazil; Hospital da Universidade Federal de Grande Dourados, Dourados, MS, Brazil; Hospital da Universidade de Brasilia, Brasilia, DF, Brazil; Hospital Universitario de Santa Maria, Santa Maria, RS, Brazil; Hospital das Clinicas da Universidade Federal de Minas Gerais, Belo Horizonte, MG, Brazil; Empresa Brasileira de Servicos Hospitalares, Brasilia, DF, Brazil

**Keywords:** primary palmar hyperhidrosis, sympathectomy, compensatory hyperhidrosis, Hyperhidrosis Disease Severity Scale, video-assisted thoracoscopic sympathectomy, unilateral sympathectomy, bilateral sympathectomy

## Abstract

**Objective:** To map, using the Hyperhidrosis Disease Severity Scale (HDSS), the preoperative distribution and six-month regional evolution of sweating severity across 20 anatomical sites after dominant-side unilateral versus one-stage bilateral R4 video-assisted thoracoscopic sympathectomy for primary palmar hyperhidrosis.

**Methods:** This was a secondary complete-case, site-level analysis of a prospective multicenter randomized trial. Adults undergoing one-stage bilateral thoracic sympathectomy (BTS) or dominant-side unilateral sympathectomy (UniS) were eligible when baseline and six-month HDSS data were complete for all 20 prespecified sites. HDSS was grouped as 1, 2, or 3-4. Regional outcomes were classified as improvement, stability, or worsening and were stratified by baseline HDSS.

**Results:** Ninety-two patients were included: 36 BTS and 56 UniS. Before surgery, severe sweating was present in 182 of 184 hand-site observations, 168 of 184 foot-site observations, and 75 of 184 axillary-site observations. At six months, improvement was more frequent after BTS than UniS in the hands (88.9% vs 52.7%) and axillae (72.2% vs 29.5%). Worsening was concentrated mainly in abdominal, hemidorsal, and thoracic sites. Among observations that were normal at baseline, progression to HDSS 3-4 occurred in 10.8% after BTS and 7.7% after UniS. Among observations with baseline HDSS 3-4, complete resolution to HDSS 1 occurred in 47.2% after BTS and 21.8% after UniS.

**Conclusion:** Baseline-stratified 20-site HDSS mapping may improve preoperative counseling and postoperative outcome assessment by distinguishing true new-onset compensatory hyperhidrosis from worsening or persistence of pre-existing sweating.

**Trial registration:** ClinicalTrials.gov NCT03921320

## Introduction

Primary hyperhidrosis is a chronic condition characterized by sweating that exceeds thermoregulatory needs and may impair social, professional, and emotional functioning (1–4). Palmar hyperhidrosis (PH) is among the most disabling presentations because hand sweating interferes with school, work, interpersonal contact, writing, device use, and other manual activities (5,6). Video-assisted thoracoscopic sympathectomy (VATS) is an established surgical option for severe palmar disease that is refractory to or unsuitable for conservative treatment (7–10).

The main limitation of thoracic sympathectomy is compensatory hyperhidrosis (CH), classically described as new sweating in areas not targeted by surgery (11–14). However, conventional postoperative questions often fail to distinguish among three clinically different situations: new sweating at a previously unaffected site, worsening of a site that was affected before surgery, and persistence of severe sweating at a site not expected to respond fully to the operation. This distinction is important because patients with PH frequently have plantar, axillary, or trunk symptoms before surgery.

The HDSS is a brief patient-reported instrument that measures the impact of sweating on daily activities and has been validated in Portuguese (15). Although the HDSS does not quantify sweat volume, it is feasible for multicenter clinical practice and allows standardized assessment of the same anatomical sites before and after surgery.

Most studies evaluating outcomes after VATS report CH as a postoperative event, often using global patient-level classifications. Postoperative sweating complaints may have different clinical meanings, however, depending on whether the affected anatomical site was normal, mildly affected, or severely affected before surgery. Sweating reported after surgery in the trunk, craniofacial region, groin, gluteal region, thighs, or feet may therefore represent true new-onset CH, worsening of pre-existing sweating, or persistence of severe sweating in a region not fully targeted by the operation.

A structured preoperative regional assessment may help distinguish these scenarios. The present secondary analysis applied the HDSS to 20 anatomical sites before and six months after R4 VATS for PH. We compared one-stage bilateral sympathectomy with dominant-side unilateral sympathectomy and examined postoperative site behavior according to baseline HDSS category. To our knowledge, this baseline-stratified regional approach has not been systematically applied to postoperative sweating after R4 VATS for PH.

### Patients and Methods

#### Study design and ethics

This was a secondary complete-case regional analysis of a prospective, multicenter, randomized comparative trial conducted in Brazil between 2019 and 2023. The source trial was approved through Plataforma Brasil (CAAE 00273818.4.1001.0068; approval decision no. 4,381,133), registered at ClinicalTrials.gov (NCT03921320), and conducted in accordance with the Declaration of Helsinki. Written informed consent was obtained from all participants. The trial protocol was published previously (16), and the primary patient-level results are available as a separate medRxiv preprint (17).

The present manuscript addresses a question distinct from the primary patient-level randomized trial report: the baseline-stratified regional evolution of sweating severity across 20 anatomical sites. The randomized allocation is described for context; because this analysis uses a complete-case subset and site-level regional observations, all inferential comparisons are interpreted as exploratory.

#### Participants and analytical cohort

Adults with PH who underwent R4 VATS during the trial were eligible. The present analysis required complete HDSS data for all 20 prespecified anatomical sites at baseline and six months. This criterion was used because incomplete regional data would prevent reliable classification of each site as improved, stable, worsened, newly affected, or persistently affected.

The regional database contained 164 operated records. Ninety-two patients met the complete-case criterion: 36 underwent one-stage bilateral thoracic sympathectomy (BTS), and 56 underwent unilateral sympathectomy on the dominant side (UniS). Missing regional data were mainly related to COVID-19 restrictions, difficulty returning for standardized follow-up, and incomplete regional fields. No imputation of missing regional HDSS values was performed.

#### Randomization and surgical strategy

In the source trial, participants were randomly assigned in a 1:1 ratio to BTS or UniS using a computer-generated allocation sequence. All operations were performed under general anesthesia by experienced teams using a standardized thoracoscopic technique. The sympathetic chain was interrupted at the fourth- rib level (R4), as specified in the published protocol (16).

In BTS, the procedure was performed bilaterally during the same anesthetic session. In UniS, only the dominant side was treated at the first operation, preserving the contralateral sympathetic chain.

#### Regional HDSS assessment and outcome definitions

Sweating severity was assessed preoperatively and six months postoperatively using the HDSS. In this analysis, HDSS 1 indicated no excessive sweating that interfered with daily activities; HDSS 2 indicated tolerable sweating with occasional interference; and HDSS 3 or 4 indicated severe sweating with frequent or constant interference. For analysis, HDSS was grouped as HDSS 1, HDSS 2, and HDSS 3-4.

Twenty prespecified anatomical sites were evaluated: right and left craniofacial, thoracic, abdominal, hemidorsal, gluteal, inguinal, thigh, foot, hand, and axillary sites. Right and left sides were analyzed as separate regional observations. Accordingly, each paired anatomical category contributed 72 observations in BTS and 112 observations in UniS.

For each site, baseline and six-month HDSS categories were compared. Improvement was defined as a lower postoperative HDSS category, stability as no change, and worsening as a higher postoperative HDSS category. Baseline-stratified analyses were performed separately for sites that were normal at baseline (HDSS 1), mildly affected (HDSS 2), or severely affected (HDSS 3-4).

True new-onset CH was defined conservatively as postoperative progression in a site with baseline HDSS 1. Progression from HDSS 1 to HDSS 2 was considered mild new postoperative sweating, whereas progression from HDSS 1 to HDSS 3-4 was considered severe new postoperative sweating. For sites with baseline HDSS 2 or HDSS 3-4, postoperative symptoms were interpreted as improvement, worsening, or persistence of pre-existing sweating rather than automatically classified as new CH.

#### Statistical analysis

Continuous variables were summarized as mean +/- standard deviation and median (interquartile range). Categorical variables were summarized as counts and percentages. Baseline characteristics were compared using Welch t tests, Mann-Whitney tests, or Fisher exact tests, as appropriate.

Postoperative HDSS distributions and regional evolution patterns were compared using Pearson chi-square tests when applicable. To reduce false-positive interpretation from multiple regional comparisons, Benjamini-Hochberg false discovery rate correction was applied within each family of anatomical-site comparisons. Because each patient contributed multiple correlated anatomical observations and this was not the primary confirmatory trial analysis, site-level p values are presented as exploratory descriptors of anatomical patterns rather than as independent confirmatory tests. Two-sided p values <0.05 were considered statistically significant before adjustment.

## Results

### Analytical cohort and baseline characteristics

Among the 92 complete-case patients, 36 underwent BTS and 56 underwent UniS. Both groups consisted predominantly of young adults, and women represented approximately two-thirds of each group. The UniS group was slightly older than the BTS group by mean age, whereas body mass index and sex distribution were similar between groups (Table 1).

**Table 1.** Baseline characteristics of complete-case patients according to surgical strategy.

| Variable | BTS (n=36) | UniS (n=56) | p value |
| --- | --- | --- | --- |
| Age (years), mean +/- SD | 24.30 +/- 5.61 | 27.00 +/- 6.83 | 0.042* |
| Age (years), median (IQR) | 22.62 (19.86-26.35) | 25.72 (20.90-31.75) | 0.053† |
| BMI (kg/m2), mean +/- SD | 23.31 +/- 3.31 | 23.27 +/- 3.00 | 0.956* |
| BMI (kg/m2), median (IQR) | 24.0 (19.75-26.0) | 24.0 (21.0-26.0) | 0.872† |
| Female sex, n (%) | 24 (66.7%) | 38 (67.9%) | 1.000‡ |
| Male sex, n (%) | 12 (33.3%) | 18 (32.1%) | 1.000‡ |

Data are presented as mean +/- standard deviation, median (interquartile range), or n (%). \*Welch t test; dagger, Mann-Whitney test; double dagger, Fisher exact test. BTS = one-stage bilateral thoracic sympathectomy; UniS = unilateral sympathectomy on the dominant side; BMI = body mass index.

### Preoperative regional distribution

Preoperative sweating severity varied substantially across anatomical categories (Table 2 and Figure 2A). Severe sweating was almost universal in the hands, affecting 182 of 184 hand-site observations (98.9%), and was also very frequent in the feet, affecting 168 of 184 foot-site observations (91.3%). Axillary involvement was heterogeneous: 54 axillary observations (29.3%) were HDSS 1, 55 (29.9%) were HDSS 2, and 75 (40.8%) were HDSS 3-4. Most craniofacial, thoracic, abdominal, hemidorsal, gluteal, inguinal, and thigh observations were HDSS 1, but a clinically meaningful proportion of these sites showed mild or severe sweating before surgery.

**Figure 1.**
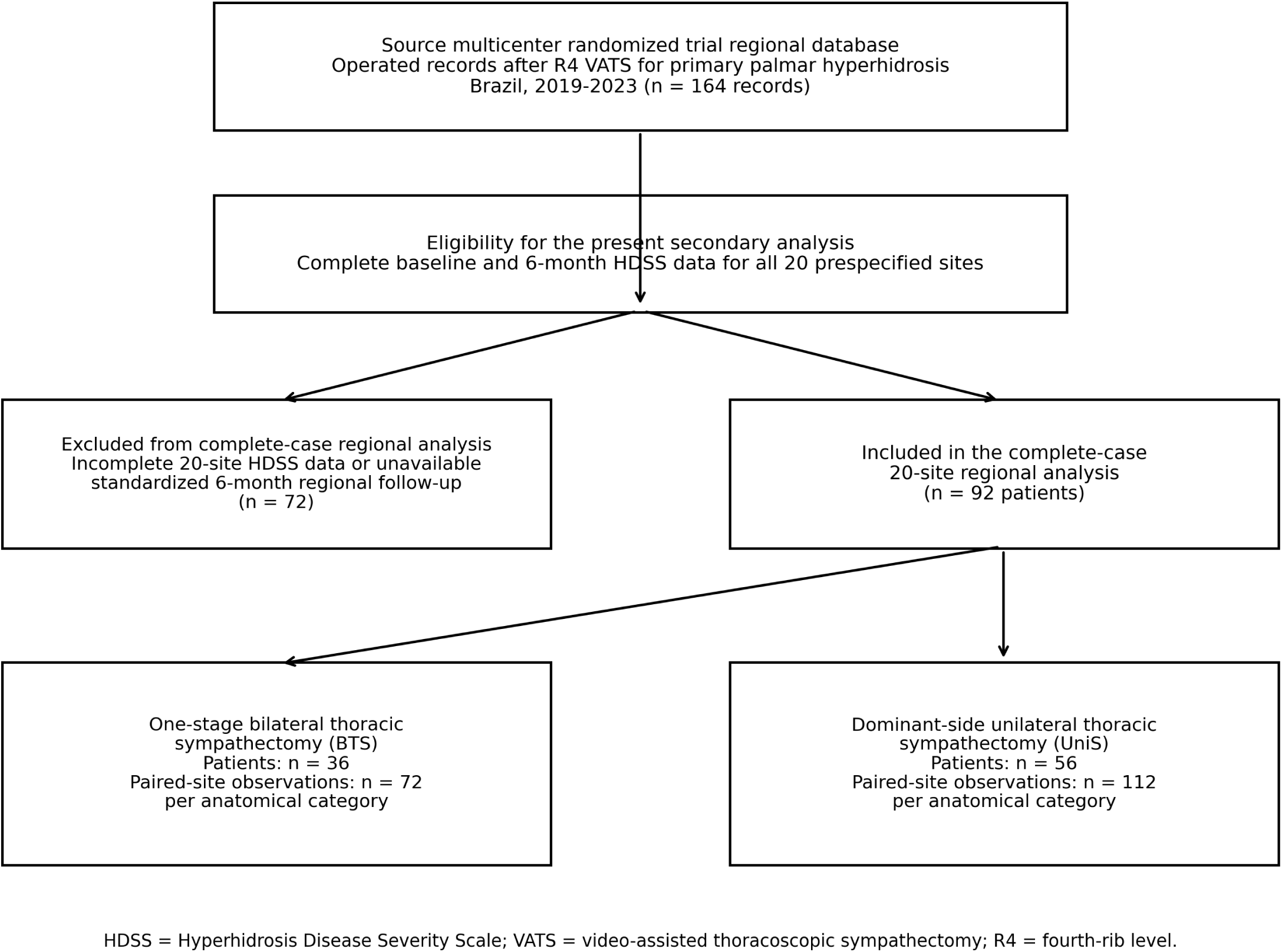
Flow diagram for the complete-case 20-site regional HDSS analysis. This figure reports the analytical cohort for the present secondary complete-case analysis and does not replace the full CONSORT diagram of the source randomized trial. The source count refers to operated records in the regional database. HDSS = Hyperhidrosis Disease Severity Scale; VATS = video-assisted thoracoscopic sympathectomy; R4 = fourth-rib level; BTS = one-stage bilateral thoracic sympathectomy; UniS = unilateral sympathectomy on the dominant side.

**Figure 2.**
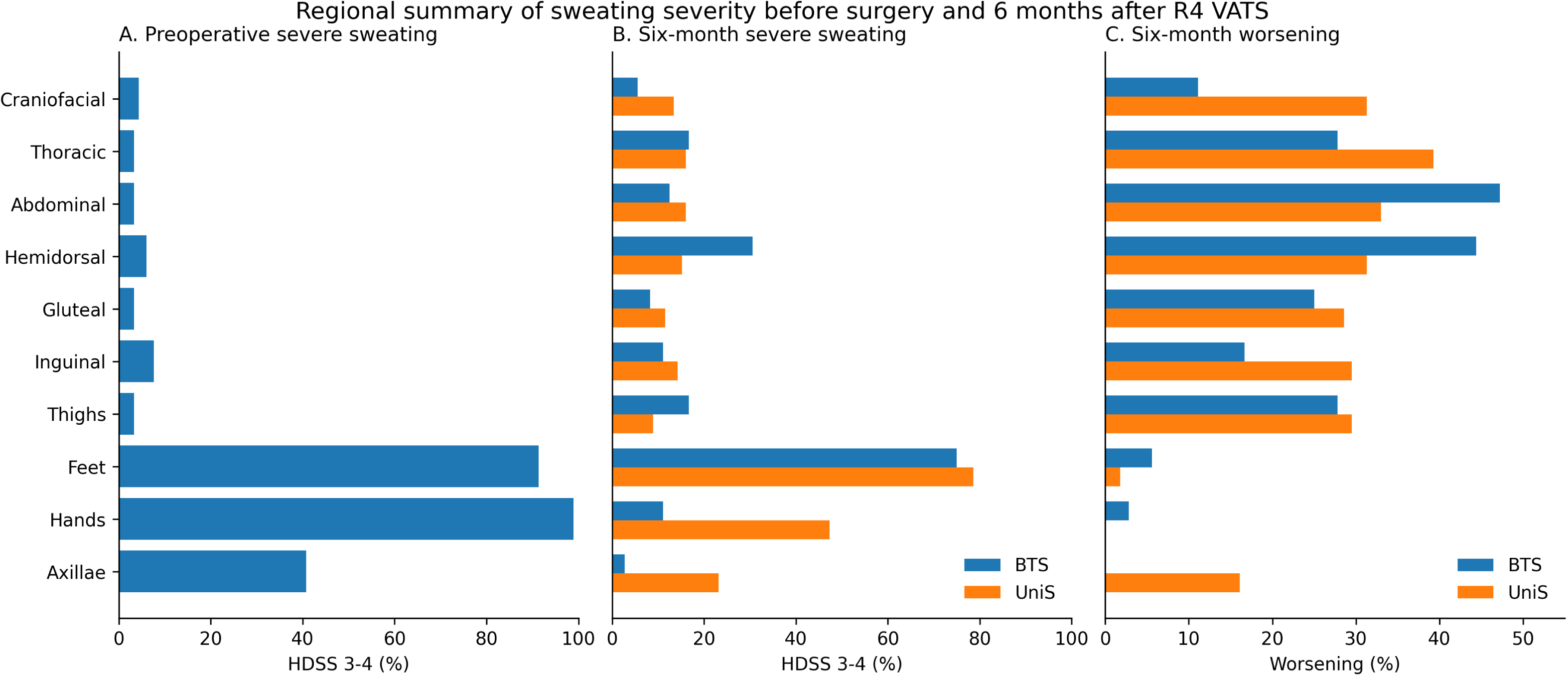
Regional summary of sweating severity before surgery and six months after R4 VATS. Panel A shows the percentage of sites with preoperative HDSS 3-4. Panel B shows six-month HDSS 3-4 by strategy. Panel C shows the percentage of sites with postoperative worsening by strategy. Anatomical categories are paired right/left sites in the regional dataset.

**Table 2.** Preoperative distribution of HDSS according to anatomical category.

| Anatomical category | HDSS 1 | HDSS 2 | HDSS 3-4 |
| --- | --- | --- | --- |
| Craniofacial | 120 (65.2%) | 56 (30.4%) | 8 (4.3%) |
| Thoracic | 129 (70.1%) | 49 (26.6%) | 6 (3.3%) |
| Abdominal | 150 (81.5%) | 28 (15.2%) | 6 (3.3%) |
| Hemidorsal | 119 (64.7%) | 54 (29.3%) | 11 (6.0%) |
| Gluteal | 142 (77.2%) | 36 (19.6%) | 6 (3.3%) |
| Inguinal | 136 (73.9%) | 34 (18.5%) | 14 (7.6%) |
| Thighs | 156 (84.8%) | 22 (12.0%) | 6 (3.3%) |
| Feet | 4 (2.2%) | 12 (6.5%) | 168 (91.3%) |
| Hands | 0 (0.0%) | 2 (1.1%) | 182 (98.9%) |
| Axillae | 54 (29.3%) | 55 (29.9%) | 75 (40.8%) |

Data are expressed as n (% of evaluated regional observations). HDSS 1 indicates absence of excessive sweating that interfered with daily activities; HDSS 2, tolerable sweating with occasional interference; and HDSS 3-4, severe sweating with frequent or constant interference.

### Six-month postoperative HDSS distribution

At six months, postoperative HDSS distribution differed most clearly in the target sites (Table 3 and Figure 2B). In the hands, BTS was associated with a greater proportion of HDSS 1 observations and fewer persistent HDSS 3-4 observations than UniS. The same pattern was observed in the axillae. After Benjamini-Hochberg correction, differences in postoperative HDSS distribution remained significant for the hands, axillae, and hemidorsal sites.

**Table 3.** Postoperative HDSS distribution at six months according to anatomical category and surgical strategy.

| Site | BTS HDSS 1 | BTS HDSS 2 | BTS HDSS 3-4 | UniS HDSS 1 | UniS HDSS 2 | UniS HDSS 3-4 | p value | BH-adjusted p |
| --- | --- | --- | --- | --- | --- | --- | --- | --- |
| Craniofacial | 48 (66.7%) | 20 (27.8%) | 4 (5.6%) | 64 (57.1%) | 33 (29.5%) | 15 (13.4%) | 0.192 | 0.320 |
| Thoracic | 42 (58.3%) | 18 (25.0%) | 12 (16.7%) | 53 (47.3%) | 41 (36.6%) | 18 (16.1%) | 0.237 | 0.339 |
| Abdominal | 32 (44.4%) | 31 (43.1%) | 9 (12.5%) | 63 (56.3%) | 31 (27.7%) | 18 (16.1%) | 0.098 | 0.196 |
| Hemidorsal | 32 (44.4%) | 18 (25.0%) | 22 (30.6%) | 47 (42.0%) | 48 (42.9%) | 17 (15.2%) | 0.012 | 0.040 |
| Gluteal | 48 (66.7%) | 18 (25.0%) | 6 (8.3%) | 64 (57.1%) | 35 (31.3%) | 13 (11.6%) | 0.427 | 0.534 |
| Inguinal | 44 (61.1%) | 20 (27.8%) | 8 (11.1%) | 63 (56.3%) | 33 (29.5%) | 16 (14.3%) | 0.756 | 0.756 |
| Thighs | 48 (66.7%) | 12 (16.7%) | 12 (16.7%) | 67 (59.8%) | 35 (31.3%) | 10 (8.9%) | 0.046 | 0.115 |
| Feet | 6 (8.3%) | 12 (16.7%) | 54 (75.0%) | 5 (4.5%) | 19 (17.0%) | 88 (78.6%) | 0.557 | 0.619 |
| Hands | 55 (76.4%) | 9 (12.5%) | 8 (11.1%) | 41 (36.6%) | 18 (16.1%) | 53 (47.3%) | 0.001 | 0.005 |
| Axillae | 55 (76.4%) | 15 (20.8%) | 2 (2.8%) | 56 (50.0%) | 30 (26.8%) | 26 (23.2%) | 0.001 | 0.005 |

Data are expressed as n (%). Pearson chi-square tests compared the distribution of HDSS 1, HDSS 2, and HDSS 3-4 between groups. Benjamini-Hochberg-adjusted p values were calculated for the 10 anatomical-category comparisons. Site-level comparisons are exploratory because multiple observations were contributed by each patient.

### Regional evolution of sweating severity

When each site was classified as improved, stable, or worsened, the response depended strongly on anatomical site and surgical strategy (Table 4 and Figure 2C). The greatest improvements occurred in the hands and axillae, especially after BTS. Hand-site improvement occurred in 88.9% of BTS observations and 52.7% of UniS observations. Axillary improvement occurred in 72.2% and 29.5%, respectively.

**Table 4.**
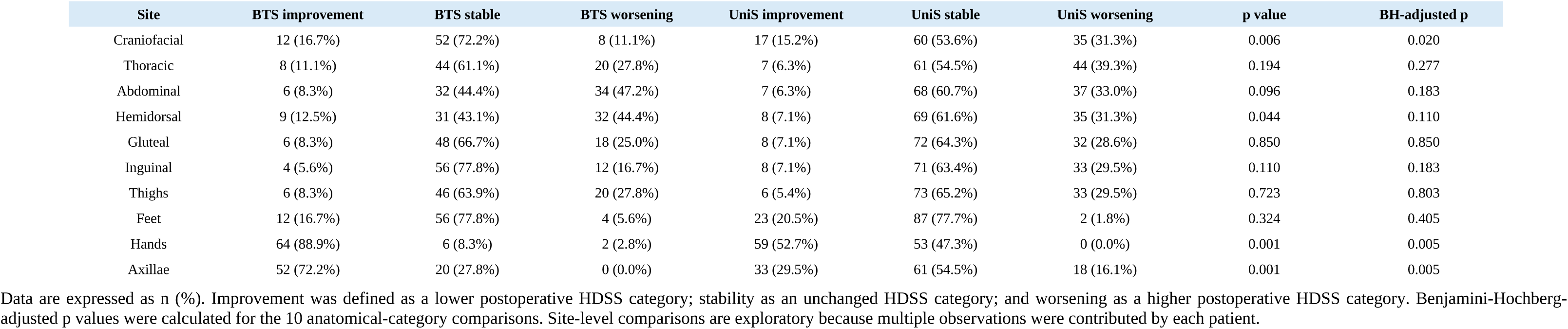
Six-month regional evolution of sweating severity according to surgical strategy.

| Site | BTS improvement | BTS stable | BTS worsening | UniS improvement | UniS stable | UniS worsening | p value | BH-adjusted p |
| --- | --- | --- | --- | --- | --- | --- | --- | --- |
| Craniofacial | 12 (16.7%) | 52 (72.2%) | 8 (11.1%) | 17 (15.2%) | 60 (53.6%) | 35 (31.3%) | 0.006 | 0.020 |
| Thoracic | 8 (11.1%) | 44 (61.1%) | 20 (27.8%) | 7 (6.3%) | 61 (54.5%) | 44 (39.3%) | 0.194 | 0.277 |
| Abdominal | 6 (8.3%) | 32 (44.4%) | 34 (47.2%) | 7 (6.3%) | 68 (60.7%) | 37 (33.0%) | 0.096 | 0.183 |
| Hemidorsal | 9 (12.5%) | 31 (43.1%) | 32 (44.4%) | 8 (7.1%) | 69 (61.6%) | 35 (31.3%) | 0.044 | 0.110 |
| Gluteal | 6 (8.3%) | 48 (66.7%) | 18 (25.0%) | 8 (7.1%) | 72 (64.3%) | 32 (28.6%) | 0.850 | 0.850 |
| Inguinal | 4 (5.6%) | 56 (77.8%) | 12 (16.7%) | 8 (7.1%) | 71 (63.4%) | 33 (29.5%) | 0.110 | 0.183 |
| Thighs | 6 (8.3%) | 46 (63.9%) | 20 (27.8%) | 6 (5.4%) | 73 (65.2%) | 33 (29.5%) | 0.723 | 0.803 |
| Feet | 12 (16.7%) | 56 (77.8%) | 4 (5.6%) | 23 (20.5%) | 87 (77.7%) | 2 (1.8%) | 0.324 | 0.405 |
| Hands | 64 (88.9%) | 6 (8.3%) | 2 (2.8%) | 59 (52.7%) | 53 (47.3%) | 0 (0.0%) | 0.001 | 0.005 |
| Axillae | 52 (72.2%) | 20 (27.8%) | 0 (0.0%) | 33 (29.5%) | 61 (54.5%) | 18 (16.1%) | 0.001 | 0.005 |

Data are expressed as n (%). Improvement was defined as a lower postoperative HDSS category; stability as an unchanged HDSS category; and worsening as a higher postoperative HDSS category. Benjamini-Hochberg-adjusted p values were calculated for the 10 anatomical-category comparisons. Site-level comparisons are exploratory because multiple observations were contributed by each patient.

Postoperative worsening was concentrated mainly in trunk and adjacent sites. The highest rates of worsening were observed in abdominal, hemidorsal, and thoracic sites. Feet showed the most stable pattern, with 77.7% unchanged and only 3.3% worsened across both strategies. After Benjamini-Hochberg correction, differences in regional evolution remained significant for craniofacial sites, hands, and axillae.

### Outcomes according to baseline HDSS severity

Baseline-stratified analysis clarified the meaning of postoperative sweating complaints. Among sites with baseline HDSS 3-4, BTS was associated with a higher rate of complete resolution to HDSS 1 than UniS (47.2% vs 21.8%) and a lower rate of persistent severe sweating (36.5% vs 60.7%). The largest differences were observed in the hands and axillae (Supplementary Table S1).

Among baseline HDSS 2 sites, postoperative behavior was heterogeneous: 40.6% of BTS observations and 29.8% of UniS observations improved, whereas 17.3% and 21.4%, respectively, worsened. Axillary sites showed the clearest benefit after BTS (Supplementary Table S2).

Among baseline HDSS 1 sites, most previously unaffected areas remained stable. However, new postoperative sweating occurred in several non-target sites. Severe new sweating, defined as progression from HDSS 1 to HDSS 3-4, occurred in 10.8% of BTS observations and 7.7% of UniS observations. This subgroup provides the closest regional representation of true CH because it isolates sites that were normal before surgery (Supplementary Table S3).

## Discussion

This secondary complete-case analysis provides a detailed regional description of the preoperative distribution and six-month evolution of sweating severity following R4 VATS for PH, including both unilateral and bilateral surgical strategies. The main originality lies not only in evaluating postoperative sweating but also in interpreting each complaint according to the baseline severity of the same anatomical site. Applying the HDSS to 20 sites before and after surgery allowed a more precise description of sites that were normal, mildly affected, or severely affected before the procedure.

This baseline-stratified approach is clinically important because postoperative sweating complaints do not represent a single phenomenon. In sites with preoperative HDSS 1, postoperative worsening is the closest clinical definition of true new-onset CH. In sites with preoperative HDSS 2, findings may represent improvement, stability, or worsening of pre-existing mild sweating. In sites with preoperative HDSS 3-4, outcomes may reflect complete resolution, partial improvement, or persistence of disabling sweating in regions that were already severely affected. The same postoperative complaint may therefore have different meanings depending on the preoperative condition of that site.

This distinction matters because CH is often discussed as a global postoperative complication without sufficient attention to the preoperative regional phenotype. Without a preoperative regional map, physicians may overestimate the incidence of true new CH and underestimate persistence or progression of pre-existing non-palmar hyperhidrosis. Postoperative sweating should be classified not only by location and severity but also by whether it appeared at a previously unaffected site, worsened at a previously affected site, or persisted despite surgery.

PH is usually presented clinically as a focal disorder (20), but the baseline distribution in this cohort demonstrates a broader sweating phenotype. Severe sweating was almost universal in the hands and very frequent in the feet, while the axillae showed mixed mild and severe involvement. Several non-palmar regions also exhibited sweating before surgery. This broader baseline phenotype helps explain why global postoperative questions about CH may be misleading.

The high prevalence of plantar sweating is particularly relevant. Severe plantar sweating was present in most foot observations at baseline, confirming that PH was frequently associated with important plantar involvement (21,22). Plantar hyperhidrosis is not the main target of R4 VATS; accordingly, the feet remained largely stable and frequently severe after surgery. Patients should be informed that successful palmar control does not necessarily imply meaningful improvement in plantar symptoms. Excellent hand outcomes may coexist with persistent disabling foot sweating, which may influence satisfaction if expectations are not addressed before surgery (22).

The use of HDSS across multiple anatomical sites was fundamental to this analysis. Objective methods such as gravimetry or evaporimetry can measure sweat production under controlled conditions, but they are difficult to apply consistently in routine multicenter surgical practice and may not capture the burden of sweating in daily life. The HDSS, although subjective and ordinal, is simple, patient-centered, and feasible. Applying the same instrument before and after surgery at each site allowed the analysis to distinguish improvement, stability, worsening of pre-existing sweating, and sweating that appeared in previously unaffected sites (15).

The surgical level helps explain the observed response. R4 VATS is expected to act most directly on upper-limb sweating, especially palmar symptoms. Consistent with this mechanism, the greatest postoperative improvements were observed in the hands and axillae. These findings support the effectiveness of an R4 strategy for the main target areas of PH (9,18). The limited effect on plantar sweating and variable response in other non-target sites highlight the heterogeneity of regional outcomes. Figure 3 summarizes the distribution before and after surgery using an HDSS-based homunculus.

**Figure 3.**
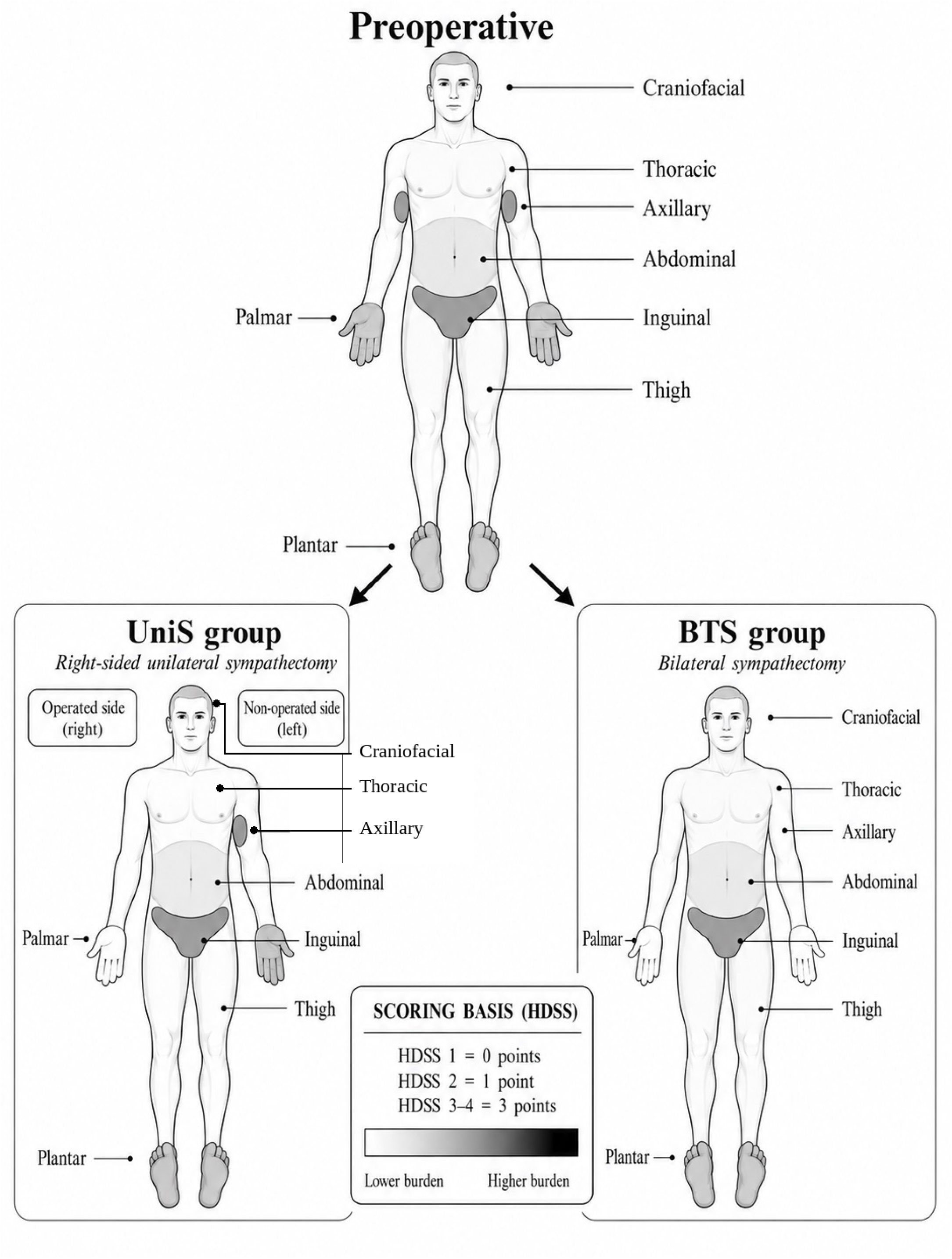
HDSS-based homunculus illustrating the regional burden of hyperhidrosis before and after R4 VATS. The lower panels summarize the unilateral (UniS) and bilateral (BTS) strategies. The UniS panel depicts a representative right-sided procedure; in the trial, UniS was performed on the dominant side. Shading is based on HDSS burden: HDSS 1 = 0 points, HDSS 2 = 1 point, and HDSS 3-4 = 3 points.

BTS produced greater improvement in the main target sites. In the hands, improvement occurred in 88.9% of site observations after BTS and 52.7% after UniS; in the axillae, improvement occurred in 72.2% and 29.5%, respectively. These differences are expected because BTS treats both sides during the same session, whereas UniS intentionally treats only the dominant side at the first operation (23). The lower apparent improvement after UniS should therefore not be interpreted as technical failure. The untreated side contributes persistent hand and axillary observations to site-level analyses.

UniS should be understood as a less extensive first-stage strategy rather than a fully equivalent bilateral operation. Its potential value lies in reducing the immediate extent of sympathetic interruption while allowing later contralateral treatment when necessary (19,23,24). The present regional results should be interpreted alongside the patient-level outcomes from the primary trial, including bilateral symptom control, quality of life, patient satisfaction, CH severity, and the need for second-stage surgery (17). Regional site-level analyses clarify anatomical patterns but do not replace evaluation of the complete patient-level surgical pathway.

Postoperative worsening was anatomically patterned rather than random. It was concentrated mainly in abdominal, hemidorsal, and thoracic sites, which are typical regions of CH after VATS (25). Craniofacial sites also changed, particularly in the unilateral group. Counseling should therefore go beyond stating that CH may occur: patients should also be told where new or worsened sweating is most likely to appear and how it differs from persistence of symptoms that were already present.

The baseline-stratified analyses refine the usual interpretation of CH. Among sites with preoperative HDSS 3-4, BTS was more effective in converting severe sweating to HDSS 1 and reducing persistent severe sweating, especially in the hands and axillae. Severe sweating in non-target areas may nevertheless persist, particularly in the feet (22). Among sites with preoperative HDSS 2, behavior was more heterogeneous and should not be interpreted using a simple improved-versus-compensatory framework. These findings complement the patient-level results of the source randomized trial (17).

Among sites with preoperative HDSS 1, postoperative worsening represented sweating emerging in previously unaffected areas and therefore corresponded most closely to classical CH (25). Even in this group, anatomical location matters. New sweating in a small or rarely noticed area may have a different impact from new trunk sweating, where clothing discomfort, visible wetness, and social embarrassment may be more relevant (26). Future studies should combine regional severity with patient-reported impact to identify the postoperative changes that are most clinically important.

Taken together, these data show that a postoperative sweating complaint may reflect at least three situations: true new sweating in a previously normal site, worsening of a site that was already affected before surgery, or persistence of severe sweating in a site not adequately influenced by the operation. Labelling all postoperative sweating as CH oversimplifies the phenomenon and may obscure the balance between benefit at target sites and burden at non-target sites. These outcomes should be described separately.

The study has several strengths. It was derived from a prospective multicenter randomized trial, used a standardized R4 surgical strategy, applied the same HDSS instrument before and after surgery, and evaluated 20 paired anatomical sites rather than a single global question. The baseline-stratified design is the main methodological contribution because it separates true new postoperative sweating from worsening or persistence of symptoms present before surgery.

Several limitations should be acknowledged. First, this was a complete-case analysis of 92 patients rather than the full randomized cohort, and selection bias cannot be excluded. Second, follow-up was limited to six months and may not capture late adaptation, progression, or persistence of compensatory symptoms. Third, the HDSS is subjective and ordinal; it captures clinical impact but not sweat volume. Fourth, each patient contributed multiple correlated regional observations, so site-level p values should be interpreted as exploratory. Fifth, quality-of-life outcomes and patient-level CH severity are not reported here because they belong to the primary randomized trial analysis. Finally, the analysis was not powered to detect small differences in anatomical sites with low baseline prevalence.

## Conclusion

In patients with PH undergoing R4 thoracoscopic sympathectomy, baseline sweating severity was distributed across multiple anatomical sites and evolved heterogeneously after surgery. BTS produced greater six-month improvement in the main target sites, especially the hands and axillae, whereas postoperative worsening was concentrated mainly in selected trunk sites. A structured baseline-stratified 20-site HDSS assessment may improve counseling and outcome assessment by distinguishing true new-onset CH from worsening or persistence of pre-existing sweating.

## Data Availability

All data produced in the present study are available upon reasonable request to the authors

## Acknowledgments

The authors acknowledge the participating centers and study teams involved in patient recruitment, surgery, and follow-up. Generative artificial intelligence was used only for language editing and manuscript formatting support. It was not used for study design, data collection, statistical analysis, data interpretation, or generation of clinical data. The authors reviewed and edited all content and take full responsibility for the manuscript.

## Data availability

The data underlying this article may be shared upon reasonable request to the corresponding author, subject to ethics approval and institutional data-sharing requirements.

## Related work

The primary patient-level randomized outcomes from the same trial are available as a separate medRxiv preprint (reference 17). The present manuscript addresses a distinct baseline-stratified site-level question and does not duplicate the primary outcome tables or figures.

## Ethics oversight

The source trial was approved through Plataforma Brasil (CAAE 00273818.4.1001.0068; approval decision no. 4,381,133), conducted in accordance with the Declaration of Helsinki, and registered at ClinicalTrials.gov (NCT03921320).

## Participant consent

Written informed consent was obtained from every participant. All necessary consent and institutional forms were archived.

## Related-work disclosure

This manuscript is a distinct secondary complete-case, site-level analysis of NCT03921320. The primary patient-level randomized outcomes are available as a separate medRxiv preprint (doi:10.64898/2026.02.18.26346562). The present manuscript addresses baseline-stratified regional evolution of sweating across 20 anatomical sites and does not duplicate the primary outcome tables or figures.

## AI disclosure

Generative artificial intelligence was used only for language editing and manuscript formatting support. It was not used for study design, data collection, statistical analysis, data interpretation, or generation of clinical data. The authors reviewed and edited all content and take full responsibility for the manuscript.

