## supplemental files for "Twenty-site HDSS mapping after thoracoscopic sympathectomy for primary palmar hyperhidrosis: a secondary complete-case analysis of a multicenter randomized trial"

Supplementary Tables S1-S3. Baseline-stratified postoperative outcomes by anatomical category.

### Supplementary Table S1. Postoperative outcomes in anatomical sites with preoperative HDSS 3-4

| Site | BTS to HDSS 1 | BTS to HDSS 2 | BTS persistent HDSS 3-4 | UniS to HDSS 1 | UniS to HDSS 2 | UniS persistent HDSS 3-4 | p value | BH-adjusted p |
|---|---|---|---|---|---|---|---|---|
| Craniofacial | 4 (66.7%) | 2 (33.3%) | 0 (0.0%) | 1 (50.0%) | 0 (0.0%) | 1 (50.0%) | 0.155 | 0.310 |
| Thoracic | 2 (50.0%) | 0 (0.0%) | 2 (50.0%) | 0 (0.0%) | 2 (100.0%) | 0 (0.0%) | 0.050 | 0.125 |
| Abdominal | 2 (100.0%) | 0 (0.0%) | 0 (0.0%) | 2 (50.0%) | 1 (25.0%) | 1 (25.0%) | 0.472 | 0.590 |
| Hemidorsal | 1 (20.0%) | 0 (0.0%) | 4 (80.0%) | 0 (0.0%) | 0 (0.0%) | 6 (100.0%) | 0.924 | 0.924 |
| Gluteal | 0 (0.0%) | 2 (100.0%) | 0 (0.0%) | 2 (50.0%) | 0 (0.0%) | 2 (50.0%) | 0.050 | 0.125 |
| Inguinal | 0 (0.0%) | 0 (0.0%) | 8 (100.0%) | 0 (0.0%) | 2 (33.3%) | 4 (66.7%) | 0.321 | 0.459 |
| Thighs | 0 (0.0%) | 2 (100.0%) | 0 (0.0%) | 0 (0.0%) | 2 (50.0%) | 2 (50.0%) | 0.759 | 0.843 |
| Feet | 4 (6.5%) | 8 (12.9%) | 50 (80.6%) | 2 (1.9%) | 18 (16.9%) | 86 (81.1%) | 0.259 | 0.432 |
| Hands | 55 (78.6%) | 9 (12.9%) | 6 (8.6%) | 41 (36.6%) | 18 (16.1%) | 53 (47.3%) | 0.001 | 0.005 |
| Axillae | 25 (69.4%) | 9 (25.0%) | 2 (5.6%) | 14 (35.9%) | 7 (17.9%) | 18 (46.2%) | 0.001 | 0.005 |
| Total | 93 (47.2%) | 32 (16.2%) | 72 (36.5%) | 62 (21.8%) | 50 (17.5%) | 173 (60.7%) | <0.001 | NA |

Data are expressed as n (% within each baseline HDSS 3-4 site and surgical group). Benjamini-Hochberg-adjusted p values were calculated for the 10 anatomical-category comparisons; the total row is descriptive and was not included in regional correction.

### Supplementary Table S2. Postoperative outcomes in anatomical sites with preoperative HDSS 2

| Site | BTS improvement | BTS stability | BTS worsening | UniS improvement | UniS stability | UniS worsening | p value | BH-adjusted p |
|---|---|---|---|---|---|---|---|---|
| Craniofacial | 6 (25.0%) | 14 (58.3%) | 4 (16.7%) | 16 (50.0%) | 12 (37.5%) | 4 (12.5%) | 0.163 | 0.293 |
| Thoracic | 6 (31.6%) | 8 (42.1%) | 5 (26.3%) | 5 (16.7%) | 13 (43.3%) | 12 (40.0%) | 0.410 | 0.461 |
| Abdominal | 4 (40.0%) | 6 (60.0%) | 0 (0.0%) | 4 (22.2%) | 10 (55.6%) | 4 (22.2%) | 0.228 | 0.342 |
| Hemidorsal | 8 (44.4%) | 4 (22.2%) | 6 (33.3%) | 8 (22.2%) | 24 (66.7%) | 4 (11.1%) | 0.008 | 0.049 |
| Gluteal | 4 (40.0%) | 4 (40.0%) | 2 (20.0%) | 6 (23.1%) | 14 (53.8%) | 6 (23.1%) | 0.591 | 0.591 |
| Inguinal | 4 (33.3%) | 8 (66.7%) | 0 (0.0%) | 6 (27.3%) | 10 (45.5%) | 6 (27.3%) | 0.133 | 0.293 |
| Thighs | 4 (50.0%) | 2 (25.0%) | 2 (25.0%) | 4 (28.6%) | 8 (57.1%) | 2 (14.3%) | 0.346 | 0.445 |
| Feet | 0 (0.0%) | 4 (66.7%) | 2 (33.3%) | 3 (50.0%) | 1 (16.7%) | 2 (33.3%) | 0.091 | 0.273 |
| Hands | 0 (0.0%) | 0 (0.0%) | 2 (100.0%) | 0 (0.0%) | 0 (0.0%) | 0 (0.0%) | NA | NA |
| Axillae | 18 (75.0%) | 6 (25.0%) | 0 (0.0%) | 12 (38.7%) | 13 (41.9%) | 6 (19.4%) | 0.011 | 0.049 |
| Total | 54 (40.6%) | 56 (42.1%) | 23 (17.3%) | 64 (29.8%) | 105 (48.8%) | 46 (21.4%) | 0.114 | NA |

Data are expressed as n (% within each baseline HDSS 2 site and surgical group). Benjamini-Hochberg-adjusted p values were calculated for applicable anatomical-category comparisons; the total row was not included in regional correction.

### Supplementary Table S3. Postoperative outcomes in anatomical sites with preoperative HDSS 1

| Site | BTS stability | BTS worsening to HDSS 2 | BTS worsening to HDSS 3-4 | UniS stability | UniS worsening to HDSS 2 | UniS worsening to HDSS 3-4 | p value | BH-adjusted p |
|---|---|---|---|---|---|---|---|---|
| Craniofacial | 38 (90.5%) | 4 (9.5%) | 0 (0.0%) | 47 (60.3%) | 21 (26.9%) | 10 (12.8%) | 0.002 | 0.016 |
| Thoracic | 34 (69.4%) | 10 (20.4%) | 5 (10.2%) | 48 (60.0%) | 26 (32.5%) | 6 (7.5%) | 0.321 | 0.367 |
| Abdominal | 26 (43.3%) | 25 (41.7%) | 9 (15.0%) | 57 (63.3%) | 27 (30.0%) | 6 (6.7%) | 0.038 | 0.101 |
| Hemidorsal | 23 (46.9%) | 14 (28.6%) | 12 (24.5%) | 39 (55.7%) | 24 (34.3%) | 7 (10.0%) | 0.105 | 0.169 |
| Gluteal | 44 (73.3%) | 12 (20.0%) | 4 (6.7%) | 56 (68.3%) | 21 (25.6%) | 5 (6.1%) | 0.736 | 0.736 |
| Inguinal | 40 (76.9%) | 12 (23.1%) | 0 (0.0%) | 57 (67.9%) | 21 (25.0%) | 6 (7.1%) | 0.127 | 0.169 |
| Thighs | 44 (71.0%) | 8 (12.9%) | 10 (16.1%) | 63 (67.7%) | 25 (26.9%) | 6 (6.5%) | 0.032 | 0.101 |
| Feet | 2 (50.0%) | 0 (0.0%) | 2 (50.0%) | 0 (0.0%) | 0 (0.0%) | 0 (0.0%) | NA | NA |
| Hands | 0 (0.0%) | 0 (0.0%) | 0 (0.0%) | 0 (0.0%) | 0 (0.0%) | 0 (0.0%) | NA | NA |
| Axillae | 12 (100.0%) | 0 (0.0%) | 0 (0.0%) | 30 (71.4%) | 10 (23.8%) | 2 (4.8%) | 0.110 | 0.169 |
| Total | 263 (67.4%) | 85 (21.8%) | 42 (10.8%) | 397 (64.0%) | 175 (28.2%) | 48 (7.7%) | 0.035 | NA |

Data are expressed as n (% within each baseline HDSS 1 site and surgical group). Stability was defined as HDSS 1 to HDSS 1; worsening to HDSS 2 as HDSS 1 to HDSS 2; and worsening to HDSS 3-4 as HDSS 1 to HDSS 3-4. Benjamini-Hochberg-adjusted p values were calculated for applicable anatomical-category comparisons; the total row was not included in regional correction.
